# Privacy Fact Sheets mitigate Disease-related Privacy Concerns and Facilitate Equal Access to the Electronic Health Record: Randomized Controlled Trial

**DOI:** 10.1101/2024.10.11.24315342

**Authors:** Niklas von Kalckreuth, Markus A. Feufel

## Abstract

**Background:** The German electronic health record (EHR) aims to enhance patient care and reduce costs, but users often worry about data security. To mitigate disease-related privacy concerns, for instance, surrounding stigmatized diseases, we test the effect of privacy fact sheets (PFS) - a concise but comprehensive transparency feature - on increasing EHR usage.

**Objective:** We investigate whether displaying a PFS shortly before upload decisions must be made mitigates disease-related privacy concerns and makes uploads more likely.

**Methods:** In an online user study, 393 German participants from the recruitment platform Prolific were asked to interact with a randomly assigned medical report that varied systematically in terms of disease-related stigma (high vs. low) and time course (acute vs. chronic). They were then asked to decide whether to upload the report to the EHR, while we systematically varied the presentation of privacy information (PFS vs. no PFS). Participants were randomly (single-blinded) assigned to one of the 8 conditions in parallel (stigma, time course, privacy information): low, acute, no PFS (52/393, 13.2%), low, chronic, no PFS (45/393, 11.5%), high, acute, no PFS (46/393, 11.7%), high, chronic, no PFS (55/393, 14%), low, acute, PFS (44/393, 11.2%), low, chronic, PFS (45/393, 10.4%), high, acute, PFS (56/393, 14.2%), high, chronic, PFS (54/393, 13.7%).

**Results:** The results show that, in general, upload behavior is negatively influenced by disease-related stigma (OR 0.130, *P*<.001) and positively influenced when a PFS is given (OR 4.527, *P*<.001). This increase was particularly pronounced for stigmatized diseases (OR 5.952, *P*=.006). Time course of diseases had no effect.

**Conclusions:** Our results demonstrate that PFSs help to increase EHR uploads by mitigating privacy concerns related to stigmatized diseases. This indicates that a PFS is mainly relevant and effective for users with increased privacy risk perceptions, while they do not hurt other users. Thus, implementing PFSs can increase the likelihood that more patients, even those with increased privacy concerns due to stigmatized diseases, upload their data to the EHR, ultimately increasing health equity. That is, PFS may help to realize EHR benefits such as more efficient healthcare processes, improved treatment outcomes, and reduced costs for more users.

**Trial Registration:** Deutsches Register Klinischer Studien DRKS00033652; https://drks.de/search/de/trial/DRKS00033652.

## Introduction

### Background

The digital connectedness between all actors in a healthcare system promises increased safety and efficiency [1–3]. The electronic health record (EHR) is one key element in this transformation process, because it allows patients’ health data (e.g., diagnoses, therapies, vaccinations, medication plans) to be readily documented, exchanged and viewed by various stakeholders [4–6]. This resulting network of care providers can make patient treatment more effective, safer, and faster across institutions [7]. For instance, pre-existing conditions, intolerances, and medication plans can be taken into account during diagnosis and treatment to prevent adverse medication interactions, duplicate diagnoses, over-treatment, and under-treatment [5]. Also, it is hoped that physicians will spend less time on obtaining patient’s medical history thanks to the EHR, which they could devote to actual patient treatment [8]. A prerequisite for the potential of EHR to realize is user engagement. In Germany, approximately 90% of residents are covered by statutory health insurance and will thus receive an EHR account by default in 2025 [9]. Yet, individuals retain the ability to opt out of using the EHR [10]. Specifically, the Patient Data Protection Act mandates that patients maintain in sole control of their data, allowing them to decide which information is stored in the EHR, who has access to it, and which data is to be deleted, which may result in missing or incomplete data [5]. Consequently, in addition the patient, physicians and other actors in the healthcare system can upload data but require the patient’s permission to do so. Also, despite its high IT-security standards, we found in previous studies that the perceived risks and benefits of using EHR is related to disease-specific privacy concerns, such as the stigma and the time course of illnesses, due to the more permanent and risky nature of the data associated with chronic conditions [11–13]. In summary, the success of the EHR hinges on patients’ actual use of the technology. In this study, we investigate the effect of privacy fact sheet (PFS) − a concise but comprehensive transparency feature − on increasing EHR usage and, specifically, to what extend the PFS can mitigate disease-related privacy concerns and increase the upload of medical reports to the EHR.

### Prior Work

“Notice and choice” is the most widely used framework for ensuring data privacy worldwide [14,15]. As its name suggests, it consists of two components: privacy notices and privacy choices. Whereas *privacy notices* explain how personal data are collected, processed, and shared with third parties, *privacy choices* give users control over various aspects of these practices, including the decision to start and terminate them [14]. Various studies indicate that, if informed by privacy notices, users are empowered to choose IT systems that match their preferences, typically those with high data security and privacy standards, and avoid less secure ones [16,17]. But the current formats used for privacy notices, most commonly *privacy policies*, tend to provide rather detailed information and often use legal jargon [18– 22], which aims to maximize legal protection of IT providers rather than to transparently inform users [23]. Research has shown that overly lengthy and complex privacy policies may ultimately serve as a ‘red flag’, leading users to loose trust in the provider, if not to discontinue technology use altogether [24,25]. Consequently, concise, easy-to-understand privacy notices are a prerequisite for users to adopt digital health technologies such as the EHR [25–27].

In contrast to privacy policies, the shorter *transparency features* have been shown to be an effective type of privacy notice, because they provide a brief and easy-to-understand overview of data privacy and security measures and are meant to inform rather than to provide legal assurance [28,29]. For instance, a study in the eCommerce domain demonstrated that displaying a transparency feature positively influences purchase numbers [29]. But increased usage does not (only) depend on the format of the privacy notice; it is also influenced by the contents it provides, including the efficacy of the mentioned data protection measures and privacy choices, and its timing, that is, when the privacy notice is given to users [30]. Our previous studies have shown that a transparency feature with a concise but comprehensive summary of all relevant contents − which we refer to as a privacy fact sheet (PFS) − positively influences EHR usage when given shortly before the upload process [31,32]. In addition, we could show that a patient-centered framing of these contents that specifies what users can do to control the EHR and emphasizes their data autonomy (e.g., you can control all of your data) has the biggest effect on EHR adoption [32].

As stated at the outset, in another line of studies we have shown that privacy concerns, intention to use the EHR and actual upload behavior are influenced by the characteristics of diseases, in particular by disease-related stigma and time course [11–13]. Disease-specific stigma has been shown to have an inhibiting influence on upload behavior and to increase the risks people perceive when they are asked to upload these diseases to the EHR [11,12]. Conversely, the time course of diseases, i.e., whether diseases are chronic rather than acute, tends to increase both privacy concerns and intention to use the EHR. That is, patients with chronic conditions recognize a greater value in using the EHR but have heightened privacy concerns when it comes to uploading chronic conditions to the EHR [13]. In this study, we aim at merging these two lines of studies to validate and extend the positive effect of a patient-framed PFS on users’ decision to upload diseases to the EHR when disease-specific privacy concerns are systematically varied.

### Aim of this Research and Approach

In this study, we test whether displaying a patient-framed PFS shortly before the decision to upload a medical report must be made increases the likelihood that users upload medical reports to the EHR for diseases that vary along two dimensions, time course and disease-specific stigma. After describing the methods and results, we discuss the implications, reflect on the study’s limitations, and conclude with a reflection on the objective of this study.

## Methods

### Ethics Approval and Consent to Participate

This study was approved by the Ethics Committee of the Department of Psychology and Ergonomics (IPA) at Technische Universität Berlin (tracking number: AWB_KAL_1_230206_Erweiterungsantrag). The study is registered as a randomized control trial at Deutsches Register Klinischer Studien (DRKS00033652). Participants volunteered to participate in the survey, and written informed consent was required to participate. On the first page of the survey, participants were told about the experimenter, the study purpose, what data were to be collected during the study, and where and for how long they would be stored. Also, participants had the possibility to download a pdf with the study information. Hence, participants were informed about the duration of the survey (approximately 8 minutes) as well as the compensation for participation.

### Participants

The online study was conducted between April 15, 2024 and May 16, 2024. Based on an a priori power analysis for a logistic regression using G*Power (version 3.1.9.7) with disease-related stigma (high vs low), time course (acute vs chronic) and privacy information (PFS vs no PFS) as binomial distributed predictors, a false positive rate α=0.05, a power of β=0.80, an estimated odds ratio for the predictor with the smallest expected effect size (time course) OR 1.7 (derived from the pre study with n=80 participants), and the probability of the outcome (upload decision) under the null hypothesis of 0.5, reflecting a conservative assumption, we aimed for a sample size of n=363 participants. To ensure this target was met, we oversampled participants by 30%, resulting in a total sample of 471 individuals. Oversampling accounted for potential exclusions due to failed attention checks, study dropouts, self-reported invalid data (approximately 20%, as indicated in preliminary studies[31,32], and prior medical histories with the diseases used in the study (approximately 10%, based on prior findings) [11]. Individuals aged 18 years and older residing in Germany were allowed to participate in the study, as the content and questions of the study were designed to fit the context of the German EHR. Another prerequisite was that participants had no personal previous experience (own illness) with the diseases mentioned in the medical reports we used for this study, as the handling of stigmatized diseases by affected persons is different from that of unaffected persons [33]. Sampling was conducted through Prolific, a crowdsourcing platform used to recruit participants for online surveys and experiments, known for its diverse participant pool and high data quality [34]. Participation was compensated with 1.78€ for 8 minutes, which corresponds to the German minimum wage. The mean value of the processing time was 8:47 minutes (SD 3:57 minutes) and the median 8:00 minutes. 471 individuals participated in the study.

### Design

We used a 2×2×2 between-subject study design with the three independent variables (IVs) stigmatization potential, time course and privacy information. Each participant was assigned to one unique combination of these conditions. As in preliminary studies, stigmatization potential (high vs low) and time course (acute vs chronic) were manipulated by displaying the diagnoses of a disease with the respective characteristics [11,12]. Additionally, privacy information (PFS vs no PFS) was manipulated by either displaying a PFS during the upload process or not. In preliminary studies, participants associate disease-related stigma with high risks [11,12], that consequences could arise in areas related to personal lifestyle, occupation, and social life if medical findings became known [12,33,35,36]. Furthermore, previous studies show that participants perceive the upload of diseases with a chronic time course as more beneficial than the upload of acute diseases [8,12,13]. Participants were randomly (single-blinded) assigned to one of the conditions in parallel (simple randomization, ratio: 1:1:1:1:1:1:1:1) using LimeSurvey’s built-in “rand” function. The dependent variable was the decision to upload the medical report, that is, whether participants were willing to upload the medical findings to the EHR [11,12,31].

### Materials

Following a common practice in technology acceptance studies [37,38], we used a case vignette to represent a typical situation in which an EHR app may be used. In particular, the case vignette depicted a situation where the participant has recently started using an EHR app and is now faced with the decision to upload a medical finding to their EHR (see Multimedia Appendix 1). Additionally, the disease/injury was described in lay terms with one to three sentences (see Multimedia Appendix 2). The stimuli used in the study were realistic but specially created for the purpose of the study. The medical reports were provided by hospitals and a medical association. To make the reports appear as realistic as possible, they were edited on the official document heads of these institutions. This was done with the permission of the institutions concerned. In selecting the diseases, both the related stigma and their time course were systematically varied. Disease-related stigma covered different risks for professional and social life, such as tests for STDs (i.e., gonorrhea and HIV) [39–42] and fractures or rheumatoid arthritis as diseases with low stigma. To reflect different time courses, diseases were divided according to an acute time course (e.g., wrist fracture and gonorrhea) and a chronic one (e.g., rheumatoid arthritis and HIV). Furthermore, diseases were selected to occur regardless of age, meaning they can affect individuals across different age groups, so that they would be perceived as realistic diseases by an age-diverse sample. Table 1 shows the diseases used as stimuli, categorized by level of perceived stigma and time course.

**Table 1.** Diseases used as stimuli, categorized by stigmatization potential (SP) and time course (TC).

| **TC**<br>**SP** | **Acute** | **Chronic** |
| --- | --- | --- |
| **Low** | Fractured wrist | Rheumatoid arthritis |
| **High** | Gonorrhea | HIV infection |

As in a previous study [12], an interactive prototype (a so-called click dummy) was used, which we created after the mobile EHR application of a German health insurance company (the BARMER) using a software for interface design (FIGMA). This prototype allows for a realistic interaction with an EHR. Specifically, the prototype gave participants the ability to upload findings, grant or revoke permissions to view findings, and create medication plans. Only the “Upload findings” function was used in this study.

We used the most effective privacy fact sheet (PFS) that we identified based on preliminary studies [31,32], which was marked by a concise but comprehensive content and a patient-centered framing, that is, a description of what the EHR allows its users to do to control their data (e.g., you can control all of your data) rather than what it does for them (e.g., the EHR keeps all of your data safe). Figure 1 shows the English version of the PFS used. The English translation of the full text can be found in the Multimedia Appendix 3.

**Figure 1.**
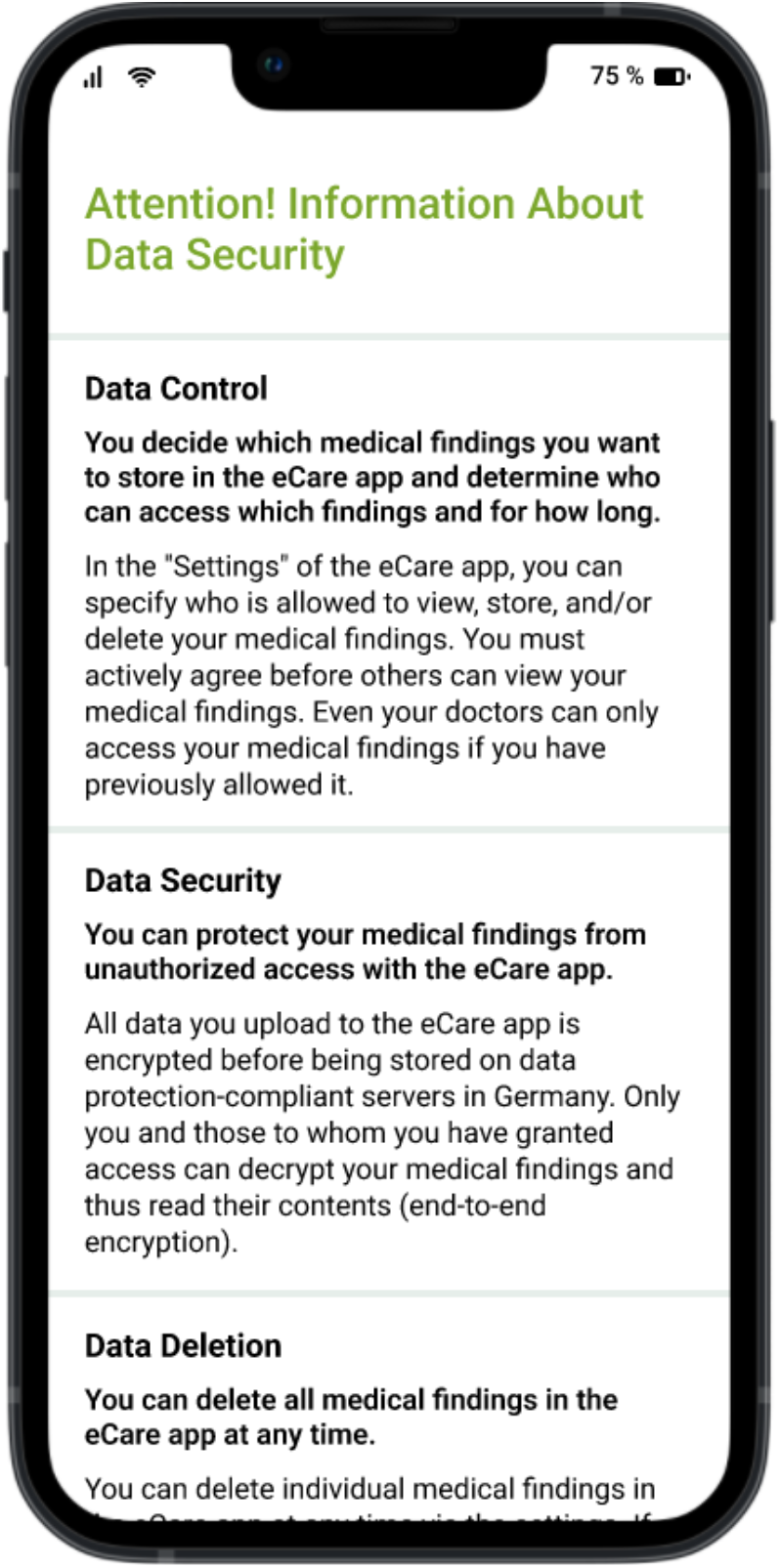
PFS used in the study.

We used LimeSurvey (version 3.28.66+230719) to create and conduct a 9-page online survey. The EHR prototype was embedded into the survey using iFrame. LimeSurvey software was used to ensure that all questions had to be answered to complete the study and receive the compensation. As in previous studies, we tested the effect of the independent variables by querying the perceived risk and perceived benefit of uploading findings to the EHR using validated items [11–13]. Also, we assumed that people perceived more risk when the stigmatization potential was high and more benefit when the time course was chronic [12,13]. Perceived risk and perceived benefit were measured using a 7-point Likert scale ranging from 1 (“Strongly disagree”) to 7 (“Strongly agree”). The decision to upload the finding was measured using a validated dichotomous item (yes/no) [11,12].

### Procedure

The study procedure is shown in Figure 2. The survey consisted of 3 parts. After giving their written informed consent according to the Declaration of Helsinki of the World Medical Association, (1) participants had several minutes to interact with the EHR prototype. (2a) Participants then read a randomly selected case vignette addressing the use of the EHR in the context of uploading a medical finding. (2b) Additionally, the participants read the medical finding of the respective illness (low or high stigmatization potential and acute or chronic time course, depending on the experimental group), as well as a brief description of the respective disease. (2c) Afterwards, as part of the upload process, the participants were asked to select the medical finding for upload. Depending on the experimental group, either a PFS was displayed before the disease could be selected or not. Participants then decided whether they wanted to upload the report to their EHR. (2d) After uploading, participants who were shown a PFS were asked a question about the content of the texts to ensure that the texts were read (attention check), and all participants were asked about the perceived privacy risks and benefits of uploading the report (manipulation check). (3) The survey was completed with the collection of demographic characteristics (age, gender, education level and experience with mHealth applications) as control variables, as well as the opportunity for participants to declare their responses invalid due to lack of care in processing them (see Multimedia Appendix 4 for the questionnaire).

**Figure 2.**
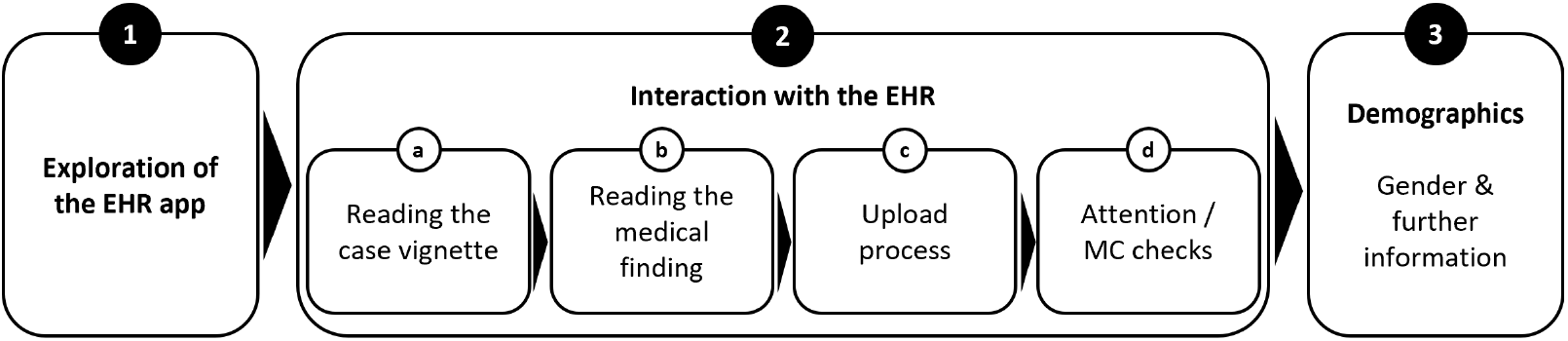
Overview of the study design.

### Hypothesis

As mentioned above, we hypothesize that diseases with high stigma would result in a high perceived risk and a chronic time course in a high perceived benefit. Hence, we hypothesize that the upload decision is negatively influenced by high disease-stigma (H1) and positively influenced by a chronic time course (H2). Based on previous studies, we also assume that a PFS will generally increase upload behavior compared to the no PFS condition (H3). More specifically regarding the aim of our study, if a PFS can mitigate disease-related concerns, we hypothesize that showing a PFS mitigates the negative influence of disease-related stigma on the upload decision (H4). Furthermore, we hypothesize that the positive influence of a chronic time course on the upload decision will be enhanced by the presence of a PFS, as it enhances perceived benefits related to long-term health management (H5). Table 2 provides an overview of the hypotheses regarding the IVs.

**Table 2.**
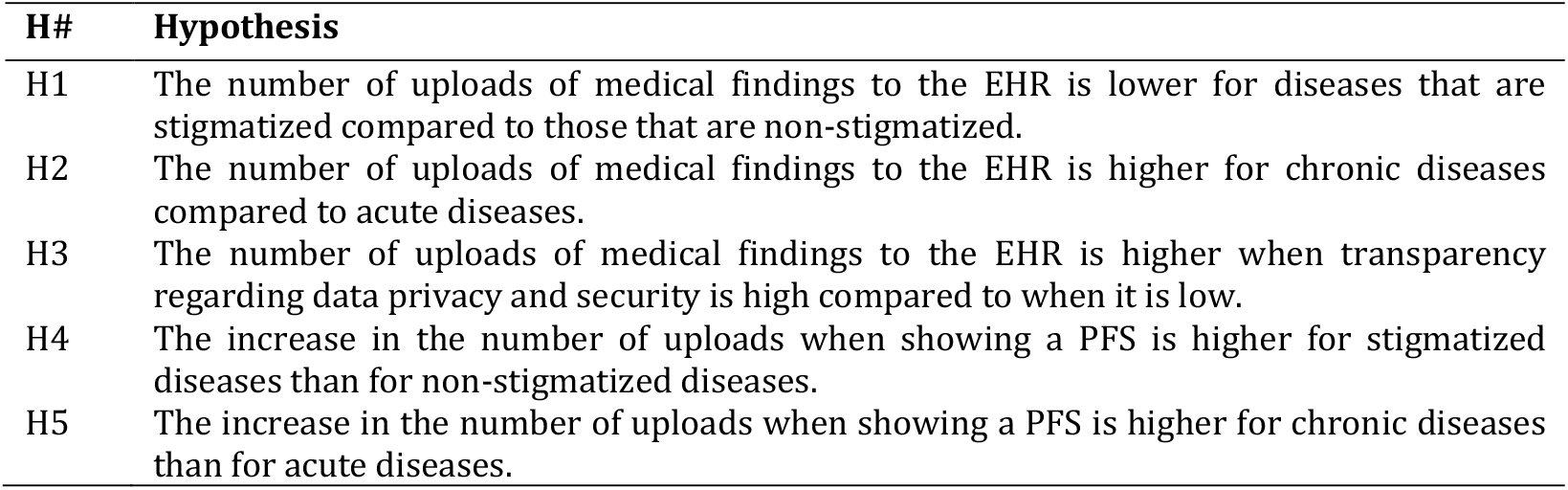
Overview of the hypotheses.

### Analyses

We cleaned and analyzed the data using RStudio (version 2023.09.1+494). The analyses regarding the manipulation checks of perceived privacy risks and benefits were performed using t-tests, a statistical method used to compare the means of two groups. The influence of the IVs (disease-specific stigma, time course, displaying a PFS) and the interaction effects between stigma and the display of a PFS, as well as between time course and the display of a PFS on the upload decision were tested using multiple logistic regression with dummy coding, a method used to model the probability of a binary outcome based on one or more predictor variables.

We also included a robustness check of the results regarding the upload decision. To control for potential influences of demographic and interindividual variables that could bias coefficients and *P* values, we used multiple logistic regression. To not bias *P* values as a result of controlling, we only included variables in the model that have been shown to have a causal relationship with the independent variables (i.e., causal confounders): age, education level, and experience with the technical system [43– 45]. *P* values were adjusted for multiple testing using the Benjamini-Hochberg procedure [46].

## Results

### Survey Characteristics

A total of 471 observations were collected. A total of 78 (16.5%) records were excluded, of which 70 (14.9%) were excluded because of incomplete questionnaires, 4 (0.85%) because participants failed the attention check, and 4 (0.85%) because responses were marked as invalid by participants. A sample of 393 observations (156 female, 231 male, 6 no information) was used for further analysis. Figure 3 shows the participation and distribution process according to the guidelines of the Consolidated Standards of Reporting Trials (CONSORT) statement [47].

**Figure 3.**
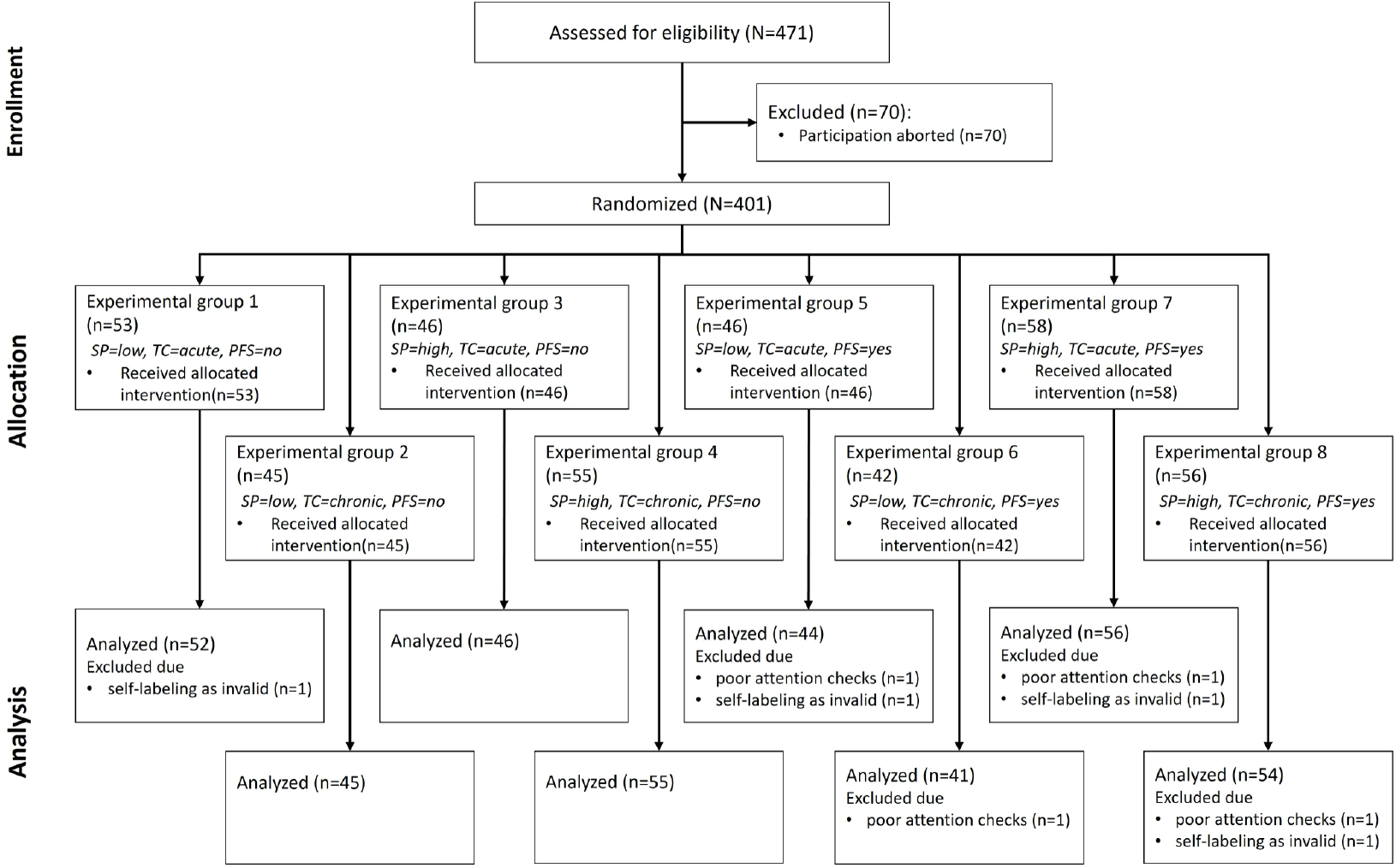
CONSORT Flow Chart. Note: SP = stigma potential, TC = time course, PFS= privacy fact sheet

Table 3 summarizes the demographic characteristics of the entire sample. The demographic characteristics of the subsamples for each experimental group are shown in the Multimedia Appendix 5.

**Table 3.** Demographic data of the sample (N=393).

| Demographic characteristic | Respondents |
| --- | --- |
| Age (years), mean (SD) | 31.67 (9.94) |
| **Gender, n (%)** |  |
| female | 156 (39.7) |
| male | 231 (58.8) |
| no answer | 6 (1.5) |
| **Education, n (%)** |  |
| No degree | 11 (2.8) |
| Highschool / vocational education | 179 (45.5) |
| Bachelor | 102 (26.0) |
| Master | 90 (22.9) |
| PhD | 11 (2.8) |
| **Experience with mHealth applications, n (%)** |  |
| No use | 226 (57.5) |
| Regular use | 167 (42.5) |

### Risk and Benefit Perception

Similar to the preliminary studies [11,12], risk and benefit perception of uploading served as a manipulation check to test the validity of our manipulation (i.e., the medical reports) with respect to the perception of risk (stigma) and benefit (time course). As expected, uploading medical findings of stigmatized diseases was perceived as riskier than those of non-stigmatized diseases (low: mean 3.88, SD 1.68; high: mean 5.15, SD 1.6; t_391_=7.648; *P*<.001). Consequently, we assume that our risk manipulation was successful. There was no significant difference of the perceived benefit regarding the time course of the disease (acute: mean 5.63, SD 1.41; chronic: mean 5.72, SD 1.16; t_391_=0.703, *P*=.483). Consequently, we assume that our benefit manipulation was not successful.

Additionally, we analyzed the benefit perception in relation to stigma and the risk perception in relation to time course, even though these were not part of the initial manipulation checks. Uploading medical findings of non-stigmatized diseases was perceived as more beneficial than those of stigmatized diseases (low: mean 5.84, SD 1.16; high: mean 5.54, SD 1.39; t_391_=2.345, *P*=.019). Furthermore, uploading medical findings of chronic diseases into the EHR was perceived as riskier than those of acute diseases (acute: mean 4.31, SD 1.83; chronic: mean 4.82, SD 1.64; t_391_=2.893, *P*=.004).

### Upload Behavior

Upload behavior was negatively associated with disease-related stigma (z=4.568, *P*<.001), thus supporting H1. Specifically, when stigma was high, it was more than seven times less likely that the report was uploaded to the EHR (76.3%; 161/211) than when stigma was low (91.8%; 167/182). Time course of the disease was not associated with the decision to upload a report (z=0.877, *P*=.380). Consequently, H2 is rejected. The PFS was positively associated with the decision to upload a medical report to the EHR (z=3.298, *P*<.001), supporting H3. When a PFS was given, participants were more than four times as likely to upload the diagnosis to their EHR (89.2%; 174/195) than when a PFS was not given (77.7%; 154/198). The absolute number of uploads is shown in Figure 5 as a function of the independent variables disease-related stigma (5A), time course (5B) and PFS/no PFS (5C).

We also tested for interaction effects between stigma and privacy information, as well as between time course and privacy information, to explore potential moderating effects. The interaction between stigma and privacy information was significant (z=2.734, *P*=.006), indicating that the increase in the number of uploads when showing a PFS is higher for stigmatized diseases than for non-stigmatized diseases, thus supporting H4 (see Figure 6A). In contrast, the interaction between time course and privacy information was not significant (z=0.094, *P*=.924), suggesting that displaying a PFS did not differentially impact the upload decision based on whether the disease was acute or chronic (see Figure 6B). Consequently, H5 is rejected. The summary of the results of the logistic regression are shown in Table 4.

**Table 4.**
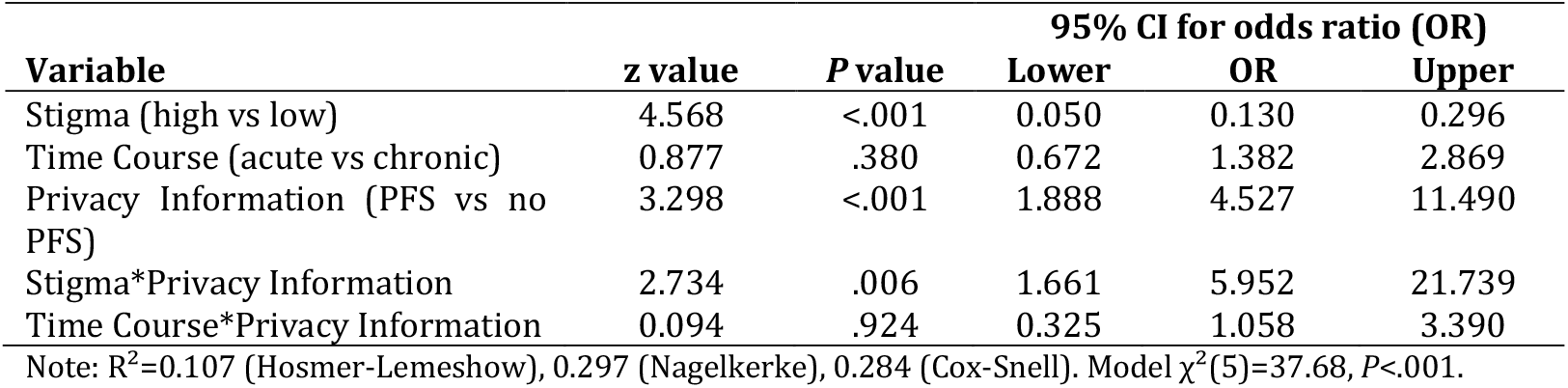
Results of the logistic regression.

| Variable | z value | P value | 95% CI for odds ratio (OR) |  |  |
| --- | --- | --- | --- | --- | --- |
|  |  |  | Lower | OR | Upper |
| Stigma (high vs low) | 4.568 | <.001 | 0.050 | 0.130 | 0.296 |
| Time Course (acute vs chronic) | 0.877 | .380 | 0.672 | 1.382 | 2.869 |
| Privacy Information (PFS vs no PFS) | 3.298 | <.001 | 1.888 | 4.527 | 11.490 |
| Stigma*Privacy Information | 2.734 | .006 | 1.661 | 5.952 | 21.739 |
| Time Course*Privacy Information | 0.094 | .924 | 0.325 | 1.058 | 3.390 |

Note: $R^2=0.107$ (Hosmer-Lemeshow), 0.297 (Nagelkerke), 0.284 (Cox-Snell). Model $\chi^2(5)=37.68$ , $P<.001$ .

### Robustness Check

When controlling for interindividual variables (age, gender, education and mHealth experience), the effects of stigma (z=4.820, *P*<.001) and information transparency (z=3.548, *P*<.001) and their interaction (z=3.086, *P*=.002) remained robust. Age had a negative effect on the upload behavior (z=2.531, *P*=.011, OR 0.965, 95% CI 0.939-0.992). With an increase in age, users were less likely to upload medical findings into their EHR. The other control variables did not influence the upload behavior.

## Discussion

### Principal Findings

The results of our study show that the decision to upload an individual medical report to the EHR is influenced by disease-related stigma as well as by privacy notices, that is, concise but comprehensive information about data privacy choices and security measures in form of PFS. As in our preliminary studies [11,12], uploading diseases with high stigma was associated with increased privacy risk perceptions than diseases with low stigma (see Figure 4A), which increased the likelihood of rejecting uploads by more than six times for stigmatized diseases compared to non-stigmatized diseases (see Figure 5A), despite a general high rating of potential benefits of uploading reports to the EHR (see Figure 4B and 4D) and the overall high willingness to upload medical findings to the EHR (see Figure 5). Furthermore, a Privacy Fact Sheet (PFS) positively influenced the decision to upload. When a PFS was displayed, the likelihood of uploading medical findings to the EHR was more than three times higher than when it was not given (see Figure 5C). This is in line with the findings of various studies showing that effective communication of data privacy choices and security information enables people to make informed decisions, thereby reducing general privacy concerns and increasing the use of EHRs [7,16,25,27,48,49].

**Figure 4.**
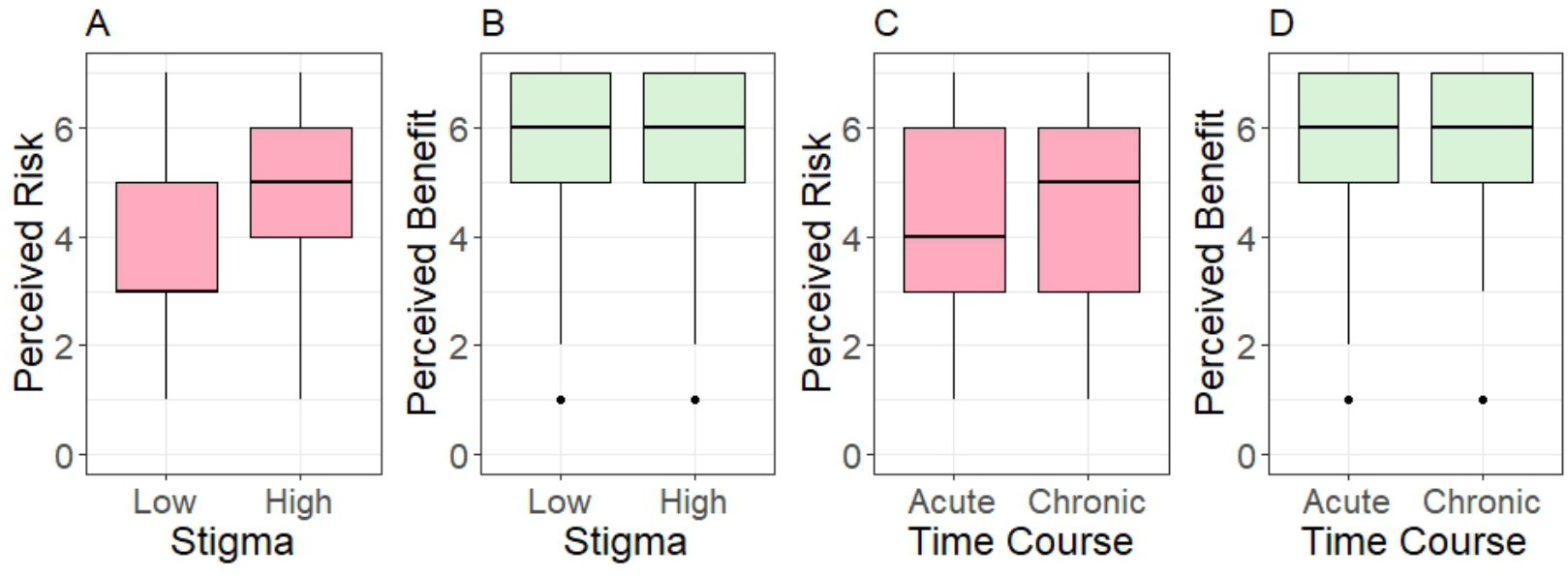
Perceived risk as a function of disease-related A) stigma and C) time course, and perceived benefit as a function of B) stigma and D) time course. The horizontal line in the box represents the median.

**Figure 5.**
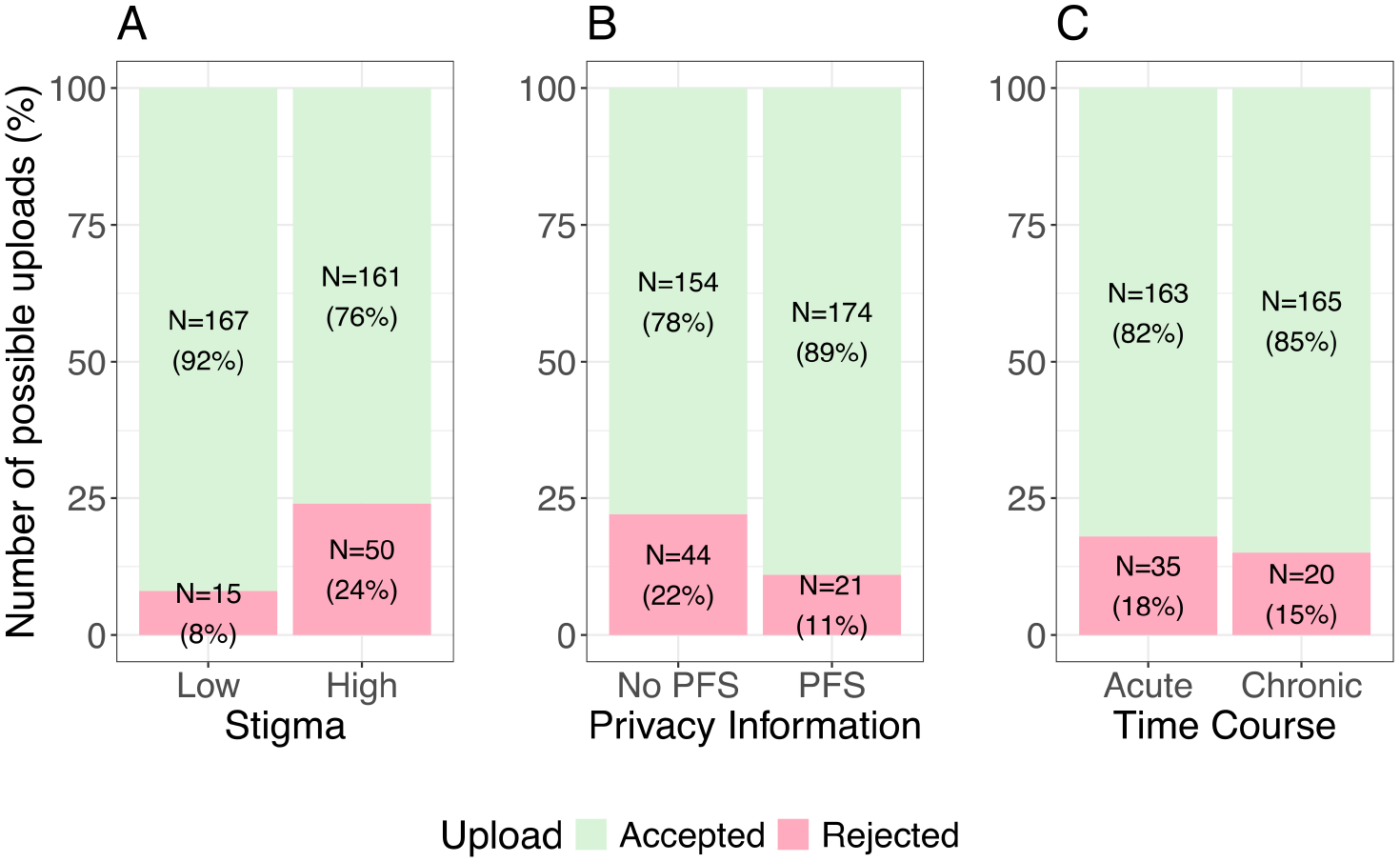
Number of uploads to the EHR as a function of disease-related (A) stigma and (B) time course and (C) privacy information.

Displaying a PFS did not influence the decision to upload medical findings for non-stigmatized diseases, as nearly all non-stigmatized medical reports were uploaded regardless of whether a PFS was given (see Figure 6A). In contrast, for stigmatized diseases, the PFS significantly increased the likelihood of uploads (see Figure 6B). This suggests that showing a PFS shortly before a decision to upload medical findings to the EHR must be made is not only effective in mitigating general privacy concerns but helps to reduce specific fears related to stigmatized diseases and increase upload decisions [28,29,31].

**Figure 6.**
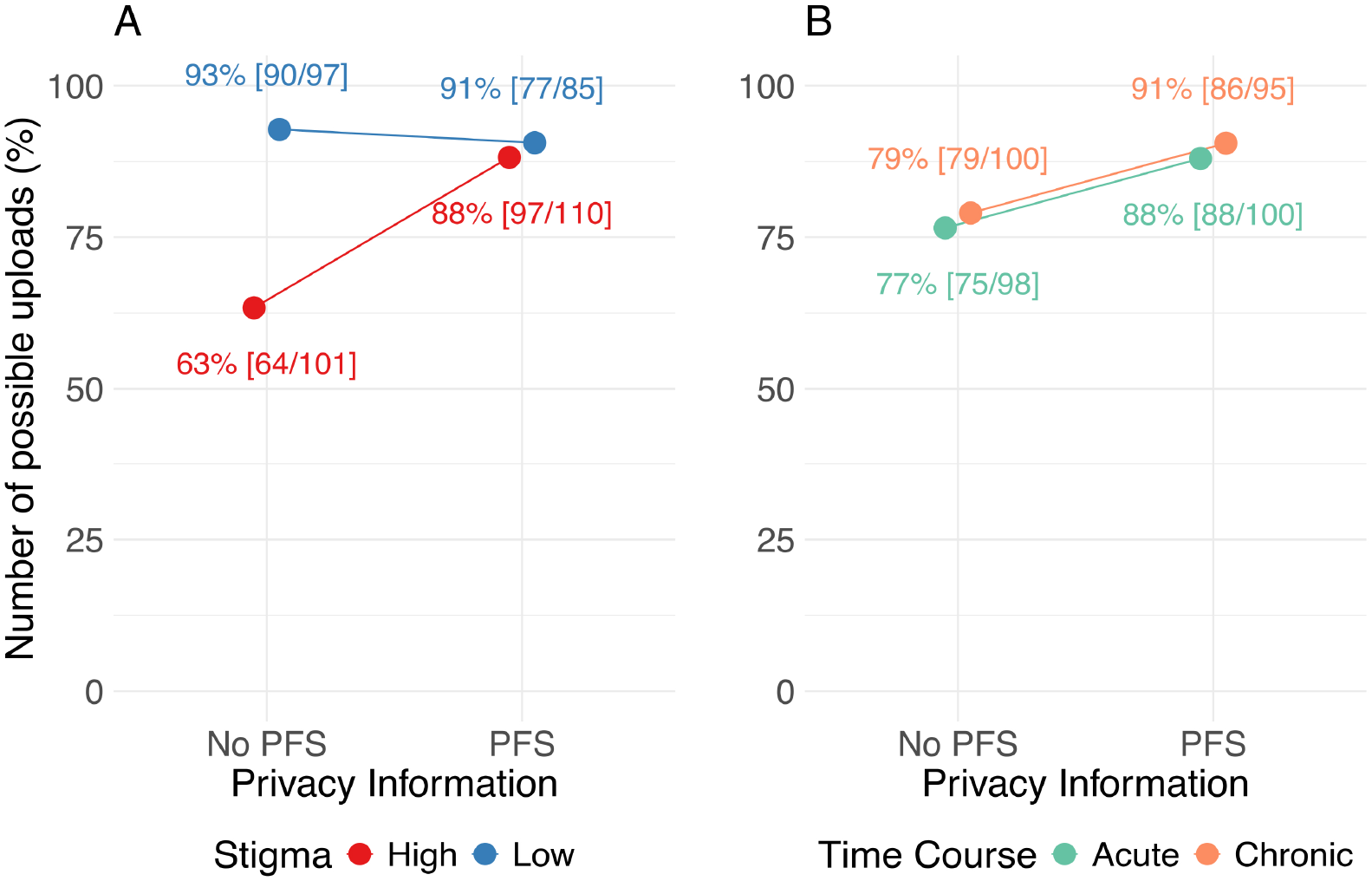
Number of uploads to the EHR as interaction between stigma and privacy information (A) and time course and privacy information (B).

More generally, studies in non-medical domains, involving low-risk scenarios such as a shopping assistant [50] and an event finder [28], showed that transparency features or the transparency of privacy policies had no effect on behavior, for instance, on the decision to access the location [50] or the intention to disclose personal data to the event finder [28]. Our findings help to explain these differing findings by showing that the relevance of transparent privacy notes is mainly contingent upon the level of perceived risk associated with the data. In low-risk scenarios, such as non-stigmatized diseases, transparency does not significantly alter behavior, as acceptance and upload rates are already high. However, in high-risk scenarios, such as those involving stigmatized health conditions, transparency features play a crucial role in mitigating concerns and can significantly enhance acceptance rates. This highlights the importance of situational context for transparency measures to matter. Transparent information about data privacy and security is not necessary in low-risk situations (although it does not hurt behavioral outcomes), but it becomes crucial for decision making in high-risk contexts, such as the handling of sensitive health data in EHRs.

### Implications

The opportunities offered by implementing transparency features in the EHR should be considered by healthcare stakeholders. Transparency features can not only reduce general privacy concerns but can also address situational concerns triggered by disease-related stigma [11]. Thus, transparency features can ultimately help to ensure equal access to EHRs, even for users who suffer from stigmatized diseases, thereby promoting health equity [35,36,51]. This way, more patients get a chance to benefit from the EHR and, as their illnesses, allergies and medications can be considered for future diagnostics and therapies, receive better and more targeted treatment.

### Limitations and Future Directions

There are several limitations in our study, which need to be considered in subsequent studies. While our manipulation checks for perceived risk (related to stigma) were successful, the manipulation of perceived benefit (related to time course) was not. This may be due to the between-subjects design of our study. In a previous within-subjects design, where participants evaluated both acute and chronic reports, the time course significantly impacted upload behavior [12]. It seems that participants, when comparing multiple conditions, can better discern when uploading is more or less beneficial. In our study, however, participants may have perceived the benefits of uploading as uniformly high, regardless of time course, leading to a diminished ability to detect differences.

It is clear that the adoption and approval of data-gathering technologies are strongly influenced by cultural differences [52]. In comparison to other European nations, the German population exhibits a heightened level of caution regarding the use of personal information online [53]. Given that in this study data collection was conducted solely with residents of Germany, future studies should validate the applicability of these findings in other countries.

We deliberately excluded participants who already had a medical history with the diseases addressed in the stimuli to avoid bias in their responses. Individuals living with a stigmatized disease are more cautious to disclose the information, especially if the disease is not immediately apparent [33,54]. The question arises to what extent the behavior of stigmatized individuals can be simulated under experimental conditions. To further strengthen the validity and generalizability of our results, a follow-up study should examine the perspective of already affected individuals.

Moreover, this was a survey study with limited immersion despite the use of an interactive click dummy. In a follow-up study, researchers could collaborate with health insurers to gather real-world data on upload behavior with a real EHR and an integrated transparency feature as used for this study. Conversely, our study faced limitations due to uncontrolled conditions like participant’s location and potential distractions, as participants completed the questionnaire online. Future research could validate our findings through a laboratory study, ensuring a more controlled environment.

Another limitation is that the distribution of our sample in terms of gender, age, and level of education does not correspond to that of the average German population [55,56]. In particular, the level of education of our sample was above average.

Although we were unable to detect any effects of the control variables gender, and level of education in the analysis, the results of this study should be validated with a more representative sample in the future.

## Conclusions

Our results show that although general upload rates to the EHR are high, disease-related stigma negatively affects upload behavior. However, displaying a transparency feature in the form of a privacy fact sheet increases the likelihood of uploads to EHRs among German adults by mitigating privacy concerns related to stigmatized diseases. Our findings indicate that the role of transparency features is contingent upon the level of perceived risk associated with the data. When the perceived risk is low, users do not need detailed privacy information to trust the technology and upload their data. However, when uploads involve sensitive data and are seen as risky, users consider privacy information and modify their upload behavior based on the information provided. Implementing transparency features in EHRs can help ensure that patients who perceive high privacy risks are not excluded from the benefits of these systems due to privacy concerns, thereby promoting health equity. This could lead to more efficient healthcare processes, improved treatment outcomes, and reduced costs within the healthcare system for more patients.

## Supporting information

Case vignettes

Short disease descriptons

Privacy Fact Sheet

Questionnaire

Demographics of all experimental groups

## Data Availability

All data produced in the present study are available upon reasonable request to the authors

## Acknowledgements

We acknowledge support from the German Research Foundation and the Open Access Publication Fund of TU Berlin. We also thank the evangelisches Studienwerk Villigst and the German Federal Ministry of Education and Research, who provided the doctoral scholarship (NvK) without which this research would not have been possible. We extend our gratitude to all participants of the study for their invaluable contributions.

## Abbreviations

CONSORT: Consolidated standards of reporting trials
EHR: Electronic health record
IoT: Internet of things
IV: Independent variable
M: Mean
mHealth: Mobile health
OR: Odds ratio
PFS: Privacy fact sheet
SD: Standard deviation
SNS: Social network site
SP: Stigmatization potential
STD: Sexually transmitted disease
TC: Time course

## References

1. Haleem A, Javaid M, Pratap Singh R, Suman R. Medical 4.0 technologies for healthcare: Features, capabilities, and applications. Internet of Things and Cyber-Physical Systems 2022;2:12–30. doi: 10.1016/j.iotcps.2022.04.001

2. Hubmann M, Pätzmann-Sietas B, Morbach H. Telemedizin und digitale Akte – Wo stehen wir?: Chancen und Herausforderungen bei der Umsetzung in Klinik-und Praxisalltag. Monatsschr Kinderheilkd 2021 Aug;169(8):711– 716. doi: 10.1007/s00112-021-01241-6

3. Kraus S, Schiavone F, Pluzhnikova A, Invernizzi AC. Digital transformation in healthcare: Analyzing the current state-of-research. Journal of Business Research 2021 Feb;123:557–567. doi: 10.1016/j.jbusres.2020.10.030

4. Bertram N, Püschner F, Gonçalves ASO, Binder S, Amelung VE. Einführung einer elektronischen Patientenakte in Deutschland vor dem Hintergrund der internationalen Erfahrungen. In: Klauber J, Geraedts M, Friedrich J, Wasem J, editors. Krankenhaus-Report 2019 Berlin, Heidelberg: Springer Berlin Heidelberg; 2019. p. 3–16. doi: 10.1007/978-3-662-58225-1_1ISBN:978-3-662-58224-4

5. Baron von Osthoff M, Watzlaw-Schmidt U, Lehmann T, Hübner J. Patientengruppenspezifische Datenhoheitsbedürfnisse und Akzeptanz der elektronischen Patientenakte. Bundesgesundheitsbl 2022 Sep 23;65:1197– 1203. doi: 10.1007/s00103-022-03589-w

6. Bundesministerium für Gesundheit. Die elektronische Patientenakte (ePA). Bundesministerium für Gesundheit. 2021. Available from: https://www.bundesgesundheitsministerium.de/elektronische-patientenakte.html [accessed Apr 14, 2023]

7. Dinev T, Albano V, Xu H, D’Atri A, Hart P. Individuals’ Attitudes Towards Electronic Health Records: A Privacy Calculus Perspective. In: Gupta A, Patel VL, Greenes RA, editors. Advances in Healthcare Informatics and Analytics Cham: Springer International Publishing; 2016. p. 19–50. doi: 10.1007/978-3-319-23294-2_2ISBN:978-3-319-23294-2

8. Kus K, Kajüter P, Arlinghaus T, Teuteberg F. Die elektronische Patientenakte als zentraler Bestandteil der digitalen Transformation im deutschen Gesundheitswesen – Eine Analyse von Akzeptanzfaktoren aus Patientensicht. HMD 2022 Dec;59(6):1577–1593. doi: 10.1365/s40702-022-00921-5

9. VDEK. Daten zum Gesundheitswesen: Versicherte. Daten zum Gesundheitswesen: Versicherte. 2024. Available from: https://www.vdek.com/presse/daten/b_versicherte.html [accessed Mar 18, 2024]

10. Bundesministerium für Gesundheit. Lauterbach: Elektr. Patientenakte ab Ende 2024 für alle verbindlich. Bundesministerium für Gesundheit. 2023. Available from: https://www.bundesgesundheitsministerium.de/presse/interviews/interview/fas-030324-elektronische-patientenakte.html [accessed Apr 14, 2023]

11. von Kalckreuth N, Feufel MA. Influence of Disease-Related Stigma on Patients’ Decisions to Upload Medical Reports to the German Electronic Health Record: Randomized Controlled Trial. JMIR Hum Factors 2024 Apr 10;11:e52625. doi: 10.2196/52625

12. von Kalckreuth N, Prümper AM, Feufel MA. The Influence of Health Data on the Use of the Electronic Health Record (EHR) – a Mixed Methods Approach. AMCIS 2023 Proceedings Panama City, Panama; 2023. Available from: https://aisel.aisnet.org/amcis2023/sig_health/sig_health/2

13. von Kalckreuth N von, Feufel MA. Disease characteristics influence the privacy calculus to adopt electronic health records: A survey study in Germany. DIGITAL HEALTH 2024;10:1–17. doi: 10.1177/20552076241274245

14. Feng Y, Yao Y, Sadeh N. A Design Space for Privacy Choices: Towards Meaningful Privacy Control in the Internet of Things. Proceedings of the 2021 CHI Conference on Human Factors in Computing Systems Yokohama, Japan: ACM; 2021. p. 1–16. doi: 10.1145/3411764.3445148

15. Sloan RH, Warner R. Beyond Notice and Choice: Privacy, Norms, and Consent. SSRN Journal 2013;14. doi: 10.2139/ssrn.2239099

16. Kelley PG, Cranor LF, Sadeh N. Privacy as part of the app decision-making process. Proceedings of the SIGCHI Conference on Human Factors in Computing Systems Paris France: ACM; 2013. p. 3393–3402. doi: 10.1145/2470654.2466466

17. Schaub F, Könings B, Weber M. Context-Adaptive Privacy: Leveraging Context Awareness to Support Privacy Decision Making. IEEE Pervasive Comput 2015 Jan;14(1):34–43. doi: 10.1109/MPRV.2015.5

18. Meier Y, Schäwel J, Krämer NC. The Shorter the Better? Effects of Privacy Policy Length on Online Privacy Decision-Making. MaC 2020 Jun 23;8(2):291–301. doi: 10.17645/mac.v8i2.2846

19. Fabian B, Ermakova T, Lentz T. Large-scale readability analysis of privacy policies. Proceedings of the International Conference on Web Intelligence Leipzig Germany: ACM; 2017. p. 18–25. doi: 10.1145/3106426.3106427

20. Mcdonald AM, Cranor LF. The Cost of Reading Privacy Policies. I/S: A Journal of Law and Policy for the Information Society 2008;4(3):543–568.

21. Reidenberg JR, Breaux T, Cranor LF, French B, Grannis A, Graves JT, Liu F, McDonald A, Norton TB, Ramanath R, Russell NC, Sadeh N, Schaub F. Disagreeable Privacy Policies: Mismatches Between Meaning and Users’ Understanding. Berkeley Technology Law Journal [University of California, Berkeley, University of California, Berkeley, School of Law]; 2015;30(1):39– 88.

22. Obar JA, Oeldorf-Hirsch A. The biggest lie on the Internet: ignoring the privacy policies and terms of service policies of social networking services. Information, Communication & Society 2020 Jan 2;23(1):128–147. doi: 10.1080/1369118X.2018.1486870

23. Braghin C, Cimato S, Della Libera A. Are mHealth Apps Secure? A Case Study. 2018 IEEE 42nd Annual Computer Software and Applications Conference (COMPSAC) Tokyo, Japan: IEEE; 2018. p. 335–340. doi: 10.1109/COMPSAC.2018.10253

24. van Haasteren A, Gille F, Fadda M, Vayena E. Development of the mHealth App Trustworthiness checklist. DIGITAL HEALTH 2019;5:1–21. doi: 10.1177/2055207619886463

25. Kulyk O, Milanovic K, Pitt J. Does My Smart Device Provider Care About My Privacy? Investigating Trust Factors and User Attitudes in IoT Systems. Proceedings of the 11th Nordic Conference on Human-Computer Interaction: Shaping Experiences, Shaping Society Tallinn Estonia: ACM; 2020. p. 1–12. doi: 10.1145/3419249.3420108

26. Krasnova H, Spiekermann S, Koroleva K, Hildebrand T. Online Social Networks: Why We Disclose. Journal of Information Technology 2010 Jun;25(2):109–125. doi: 10.1057/jit.2010.6

27. Schaub F, Balebako R, Durity AL, Cranor LF. A Design Space for Effective Privacy Notices*. In: Selinger E, Polonetsky J, Tene O, editors. The Cambridge Handbook of Consumer Privacy 1st ed Cambridge University Press; 2018. p. 365–393. doi: 10.1017/9781316831960.021ISBN:978-1-316-83196-0

28. Karwatzki S, Dytynko O, Trenz M, Veit D. Beyond the Personalization– Privacy Paradox: Privacy Valuation, Transparency Features, and Service Personalization. Journal of Management Information Systems 2017 Apr 3;34(2):369–400. doi: 10.1080/07421222.2017.1334467

29. Tsai JY, Egelman S, Cranor L, Acquisti A. The Effect of Online Privacy Information on Purchasing Behavior: An Experimental Study. Information Systems Research 2011 Jun;22(2):254–268. doi: 10.1287/isre.1090.0260

30. Adjerid I, Acquisti A, Brandimarte L, Loewenstein G. Sleights of privacy: framing, disclosures, and the limits of transparency. Proceedings of the Ninth Symposium on Usable Privacy and Security Newcastle United Kingdom: ACM; 2013. p. 1–11. doi: 10.1145/2501604.2501613

31. von Kalckreuth N, Kopka M, Appel J, Feufel MA. Unlocking the potential of the electronic health record - the influence of transparency features. ECIS 2024 Proceedings 2024. Available from: https://aisel.aisnet.org/ecis2024/track18_healthit/track18_healthit/5

32. von Kalckreuth N von, Feufel MA. Enhancing Uploads of Health Data in the Electronic Health Record - The Role of Framing and Length of Privacy Information. medRxiv; 2024. doi: 10.1101/2024.10.08.24315097

33. Goffman E. Stigma: Notes on the Management of Spoiled Identity. Englewood Cliffs, NJ: Prentice-Hall, Inc.; 1963.

34. Peer E, Brandimarte L, Samat S, Acquisti A. Beyond the Turk: Alternative platforms for crowdsourcing behavioral research. Journal of Experimental Social Psychology 2017 May;70:153–163. doi: 10.1016/j.jesp.2017.01.006

35. Stablein T, Hall JL, Pervis C, Anthony DL. Negotiating stigma in health care: disclosure and the role of electronic health records. Health Sociology Review 2015 Sep 2;24(3):227–241. doi: 10.1080/14461242.2015.1078218

36. Link BG, Phelan JC. Conceptualizing Stigma. Annu Rev Sociol 2001 Aug;27(1):363–385. doi: 10.1146/annurev.soc.27.1.363

37. Cheng Y-H, Yeh Y-J. Exploring radio frequency identification technology’s application in international distribution centers and adoption rate forecasting. Technological Forecasting and Social Change 2011 May;78(4):661–673. doi: 10.1016/j.techfore.2010.10.003

38. Miltgen CL, Popovič A, Oliveira T. Determinants of end-user acceptance of biometrics: Integrating the “Big 3” of technology acceptance with privacy context. Decision Support Systems 2013 Dec;56:103–114. doi: 10.1016/j.dss.2013.05.010

39. Bickford J, Barton SE, Mandalia S. Chronic genital herpes and disclosure…. The influence of stigma. Int J STD AIDS 2007 Sep 1;18(9):589– 592. doi: 10.1258/095646207781568484

40. Zacks S, Beavers K, Theodore D, Dougherty K, Batey B, Shumaker J, Galanko J, Shrestha R, Fried MW. Social Stigmatization and Hepatitis C Virus Infection:Journal of Clinical Gastroenterology 2006 Mar;40(3):220–224. doi: 10.1097/00004836-200603000-00009

41. Berger BE, Ferrans CE, Lashley FR. Measuring stigma in people with HIV: Psychometric assessment of the HIV stigma scale. Res Nurs Health 2001 Dec;24(6):518–529. doi: 10.1002/nur.10011

42. Zarei N, Joulaei H, Darabi E. Stigmatized Attitude of Healthcare Providers: A Barrier for Delivering Health Services to HIV Positive Patients. IJCBNM 2015;3(4):292–300.

43. Schomakers E-M, Lidynia C, Vervier LS, Calero Valdez A, Ziefle M. Applying an Extended UTAUT2 Model to Explain User Acceptance of Lifestyle and Therapy Mobile Health Apps: Survey Study. JMIR Mhealth Uhealth 2022 Jan 18;10(1):1–16. doi: 10.2196/27095

44. Uncovska M, Freitag B, Meister S, Fehring L. Patient Acceptance of Prescribed and Fully Reimbursed mHealth Apps in Germany: An UTAUT2-based Online Survey Study. J Med Syst 2023 Jan 27;47(1):1–14. doi: 10.1007/s10916-023-01910-x

45. von Kalckreuth N, Feufel MA. Extending the Privacy Calculus to the mHealth Domain: Survey Study on the Intention to Use mHealth Apps in Germany. JMIR Hum Factors 2023 Aug 16;10:e52625. doi: 10.2196/45503

46. Benjamini Y, Hochberg Y. Controlling the False Discovery Rate: A Practical and Powerful Approach to Multiple Testing. Journal of the Royal Statistical Society: Series B (Methodological) 1995 Jan;57(1):289–300. doi: 10.1111/j.2517-6161.1995.tb02031.x

47. Cuschieri S. The CONSORT statement. Saudi J Anaesth 2019;13(5):27– 30. doi: 10.4103/sja.SJA_559_18

48. Cherif E, Bezaz N, Mzoughi M. Do personal health concerns and trust in healthcare providers mitigate privacy concerns? Effects on patients’ intention to share personal health data on electronic health records. Social Science & Medicine 2021 Aug;283:114146. doi: 10.1016/j.socscimed.2021.114146

49. Kroll T, Stieglitz S. Digital nudging and privacy: improving decisions about self-disclosure in social networks. Behaviour & Information Technology 2021 Jan 2;40(1):1–19. doi: 10.1080/0144929X.2019.1584644

50. Betzing JH, Tietz M, vom Brocke J, Becker J. The impact of transparency on mobile privacy decision making. Electronic Markets 2019;30(3):607–625. doi: 10.1007/s12525-019-00332-3

51. Hatzenbuehler ML, Phelan JC, Link BG. Stigma as a Fundamental Cause of Population Health Inequalities. Am J Public Health 2013 May;103(5):813– 821. doi: 10.2105/AJPH.2012.301069

52. Krasnova H, Veltri NF, Günther O. Self-disclosure and Privacy Calculus on Social Networking Sites: The Role of Culture: Intercultural Dynamics of Privacy Calculus. Bus Inf Syst Eng 2012 Jun;4(3):127–135. doi: 10.1007/s12599-012-0216-6

53. Custers B, Dechesne F, Sears AM, Tani T, van der Hof S. A comparison of data protection legislation and policies across the EU. Computer Law & Security Review 2018 Apr;34(2):234–243. doi: 10.1016/j.clsr.2017.09.001

54. Weiss MG, Ramakrishna J, Somma D. Health-related stigma: Rethinking concepts and interventions. Psychology, Health & Medicine 2006 Aug;11(3):277–287. doi: 10.1080/13548500600595053

55. Statistisches Bundesamt. Bildungsstand. Statistisches Bundesamt. 2020. Available from: https://www.destatis.de/DE/Themen/Gesellschaft-Umwelt/Bildung-Forschung-Kultur/Bildungsstand/_inhalt.html [accessed Aug 19, 2024]

56. Statistisches Bundesamt. Bevölkerungsstand. Statistisches Bundesamt. 2022. Available from: https://www.destatis.de/DE/Themen/Gesellschaft-Umwelt/Bevoelkerung/Bevoelkerungsstand/_inhalt.html [accessed Aug 19, 2024]

