## Supplementary material for "Privacy Fact Sheets mitigate Disease-related Privacy Concerns and Facilitate Equal Access to the Electronic Health Record: Randomized Controlled Trial": Case vignettes

**Case vignette 1 (low stigma, acute time course)**
Imagine the following scenario:

1. You have recently started using your health insurance's electronic health record (EHR), the 'eCare' app, to manage your medical data and documents and share them with your doctors upon request.
2. You recently broke your wrist, but it has now fully healed.
3. You are now faced with the decision of whether or not to include the medical report of this condition in your EHR.

**Case vignette 2 (low stigma, chronic time course)**
Imagine the following scenario:

1. You have recently started using your health insurance's electronic health record (EHR), the 'eCare' app, to manage your medical data and documents and share them with your doctors upon request.
2. You have been suffering from rheumatoid arthritis for several years.
3. You are now faced with the decision of whether or not to include the medical report of this condition in your EHR.

**Case vignette 3 (high stigma, acute time course)**
Imagine the following scenario:

1. You have recently started using your health insurance's electronic health record (EHR), the 'eCare' app, to manage your medical data and documents and share them with your doctors upon request.
2. You recently became infected with a sexually transmitted disease (gonorrhea), but it has now fully healed.
3. You are now faced with the decision of whether or not to include the medical report of this condition in your EHR.

**Case vignette 4 (high stigma, chronic time course)**
Imagine the following scenario:

1. You have recently started using your health insurance's electronic health record (EHR), the 'eCare' app, to manage your medical data and documents and share them with your doctors upon request.
2. You have recently become infected with HIV.
3. You are now faced with the decision of whether or not to include the medical report of this condition in your EHR.
