## Supplementary material for "Privacy Fact Sheets mitigate Disease-related Privacy Concerns and Facilitate Equal Access to the Electronic Health Record: Randomized Controlled Trial": Short disease descriptons

### Description of the diseases

**Gonorrhea**
Gonorrhea (gonorrhea) is an infectious disease that is mainly transmitted during sex. The pathogens are bacteria known as gonococci. Gonorrhea is one of the most common sexually transmitted diseases, also known as STDs. Gonorrhea can usually be treated well with antibiotics. If it remains untreated, complications are possible - such as joint inflammation, abdominal pain or fertility problems.

**Depression**
Depression is a mental illness that can manifest itself in numerous symptoms. A persistently depressed mood, inhibition of drive and thinking, loss of interest and a variety of physical symptoms, ranging from insomnia to appetite disorders and pain, are possible signs of depression. The majority of those affected have suicidal thoughts sooner or later, and 10 to 15% of all patients with recurring severe depressive phases die by suicide. Once the correct diagnosis has been made, the situation is anything but hopeless. In recent decades, a lot has been done in terms of treatment and more than 80% of sufferers can be helped permanently and successfully.

**Wrist fracture**
A wrist fracture is a fracture of the radius (one of the two forearm bones) close to the wrist. The medical term is "distal radius fracture". Wrist fractures are the most common form of bone fracture in adults. A wrist fracture often heals without any problems, especially in the case of stable fractures. In very rare cases, complications and late effects develop, such as limited mobility or loss of strength in the wrist and fingers.

**Diabetes**
Type 1 diabetes is an autoimmune disease in which the patient's own immune system attacks the body's own insulin production in the pancreas and destroys the insulin-producing cells: this results in an absolute insulin deficiency, which leads to a sharp rise in blood sugar and a simultaneous undersupply of the body's cells. Lifelong therapy with insulin injections is therefore necessary, as well as adapting the diet to the insulin dosage in order to prevent blood sugar fluctuations. Accompanying symptoms are often other autoimmune diseases.
