## Supplementary material for "Privacy Fact Sheets mitigate Disease-related Privacy Concerns and Facilitate Equal Access to the Electronic Health Record: Randomized Controlled Trial": Privacy Fact Sheet

### Attention! Information About Data Security

#### Data Control

**You decide which medical findings you want to store in the eCare app and determine who can access which findings and for how long.**

In the "Settings" of the eCare app, you can specify who is allowed to view, store, and/or delete your medical findings. You must actively agree before others can view your medical findings. Even your doctors can only access your medical findings if you have previously allowed it.

#### Data Security

**You can protect your medical findings from unauthorized access with the eCare app.**

All data you upload to the eCare app is encrypted before being stored on data protection-compliant servers in Germany. Only you and those to whom you have granted access can decrypt your medical findings and thus read their contents (end-to-end encryption).

#### Data Deletion

**You can delete all medical findings in the eCare app at any time.**

You can delete individual medical findings in the eCare app at any time via the settings. If you no longer wish to use eCare at all, you can completely delete your eCare account and all your medical findings. Simply revoke your consent to the privacy and terms of use by email to.
