## Supplementary material for "Privacy Fact Sheets mitigate Disease-related Privacy Concerns and Facilitate Equal Access to the Electronic Health Record: Randomized Controlled Trial": Questionnaire

**Manipulation check (perceived risk):** Rate the following statement: If the information in this finding fell into the wrong hands, it would do me a lot of damage. (von Kalckreuth & Feufel, 2023)

**Manipulation check (perceived benefit):** Rate the following statement: I think it is very beneficial that physicians who treat me have access to this finding through the electronic health record (EHR). (von Kalckreuth & Feufel, 2023)

**Attention check**: Please answer the following question regarding the content of the privacy information displayed in the click dummy: At what point can the stored records be deleted? (Query as multiple choice) (self-constructed)

**Behavioral decision:** Would you like to upload the report to your electronic health record? (y/n) (von Kalckreuth et al., 2023)

### Demographics

**Age**: Please enter your age.

**Gender (m/f/d):** Please indicate your gender.

**Education**: Please enter your highest qualification.

- No degree
- School leaving certificate
- Secondary school certificate
- General qualification for university entrance
- Vocational training
- University degree (bachelor's or master's)
- other

**Experience with mHealth apps**: How often do you use health or fitness apps?

- Never
- Tried once
- One app regularly
- Multiple apps regularly

**Reability Check:** Is there any reason why we should NOT use your data? You will be paid regardless of your response.

- Yes, I rushed through.
- Yes, I did not really read the questions.
- Yes, I choose random answers.
- Yes, for other reasons.
- No, you can use my data.
