## Supplementary material for "Privacy Fact Sheets mitigate Disease-related Privacy Concerns and Facilitate Equal Access to the Electronic Health Record: Randomized Controlled Trial": Demographics of all experimental groups

*Table 1. Demographic data of the sample regarding all experimental groups (N=393). Note: SP = stigma potential, TC = time course, PFS= privacy fact sheet*

| **Demographic characteristic** | **Low SP, acute TC, no PFS (n=52)** | **Low SP, chronic TC, no PFS (n=45)** | **High SP, acute TC, no PFS (n=46)** | **High SP, chronic TC, no PFS (n=55)** | **Low SP, acute TC, with PFS (n=44)** | **Low SP, chronic TC, with PFS (n=41)** | **High SP, acute TC, with PFS (n=56)** | **High SP, chronic TC, with PFS (n=54)** | **Total (N=393)** |
|---|---|---|---|---|---|---|---|---|---|
| Age (years), mean (SD) | 30.04 (7.18) | 32.18 (11.14) | 31.11 (11.09) | 33.04 (10.68) | 30.50 (9.62) | 31.07 (9.09) | 31.54 (9.41) | 32.72 (10.11) | 31.67 (9.94) |
| **Gender, n (%)** | | | | | | | | | |
| female | 20 (38.5) | 22 (48.9) | 18 (39.1) | 26 (47.3) | 15 (34.1) | 15 (36.6) | 19 (33.9) | 21 (38.9) | 156 (39.7) |
| male | 30 (57.7) | 22 (48.9) | 27 (58.7) | 29 (52.7) | 28 (63.6) | 25 (61.0) | 37 (66.1) | 33 (61.1) | 231 (58.8) |
| no answer | 2 (3.8) | 1 (2.2) | 1 (2.2) | 0 (0) | 1 (2.3) | 1 (2.4) | 0 (0) | 0 (0) | 6 (1.5) |
| **Education, n (%)** | | | | | | | | | |
| No degree | 0 (0) | 2 (4.4) | 0 (0) | 4 (7.3) | 1 (2.3) | 2 (4.9) | 1 (1.8) | 1 (1.9) | 11 (2.8) |
| Highschool / vocational education | 29 (55.8) | 17 (37.8) | 20 (43.5) | 27 (49.1) | 20 (45.5) | 19 (46.3) | 24 (42.9) | 23 (42.6) | 179 (45.5) |
| Bachelor | 12 (23.1) | 12 (26.7) | 12 (26.1) | 15 (27.3) | 12 (27.3) | 12 (29.3) | 15 (26.8) | 12 (22.2) | 102 (26.0) |
| Master | 10 (19.2) | 8 (17.8) | 14 (30.4) | 9 (16.4) | 9 (20.5) | 8 (19.5) | 14 (25.0) | 18 (33.3) | 90 (22.9) |
| PhD | 1 (1.9) | 6 (13.3) | 0 (0) | 0 (0) | 2 (4.5) | 0 (0) | 2 (3.6) | 0 (0) | 11 (2.8) |
| **Experience with mHealth applications, n (%)** | | | | | | | | | |
| No use | 26 (50.0) | 29 (64.4) | 27 (58.7) | 34 (61.8) | 24 (54.5) | 24 (58.5) | 28 (50.0) | 36 (66.7) | 226 (57.5) |
| Regular use | 26 (50.0) | 16 (35.6) | 27 (41.3) | 21 (38.2) | 20 (45.5) | 17 (41.5) | 28 (50.0) | 18 (33.3) | 167 (42.5) |
